# Chrononutrition and prenatal mental health: The relationship between food intake indicators and prenatal depressive symptomatology

**DOI:** 10.64898/2026.08.19.26360839

**Authors:** Christina M. Personette, David Phan, Daisy Duan, Namhyun Kim, Kaleab Z. Abebe, Christina M. Scifres, Tina Costacou, Patrick Catalano, Hyagriv Simhan, Esa M Davis, Dara Mendez, Marquis S. Hawkins

## Abstract

**Aim:** Examine cross-sectional associations between mid-pregnancy food intake indicators and prenatal depressive symptomatology.

**Methods:** This secondary analysis of the Comparison of Two Screening Strategies for Gestational Diabetes trial (N = 718) examined domains of mid-pregnancy food intake (direct timing, energy timing, meal/snack structure, meal energy distribution, diet quality) derived from 24-hour dietary recalls. Depressive symptoms were measured with the Edinburgh Postnatal Depression Scale (EPDS). Generalized linear models examined associations between food intake indicators, total, and high (EPDS ≥13) depressive symptoms.

**Results:** Mean (SD) EPDS score was [6.3 (4.9)]; 12.4% (n = 89) had high depressive symptoms. Eating frequency (B = 0.08 [0.02, 0.14], p = 0.009), snack frequency (B = 0.06 [0.00, 0.12], p = 0.040), nighttime snacking frequency (B = 0.06 [0.00, 0.11], p = 0.041), and total daily energy intake (B = 0.06 [0.01, 0.12], p = 0.031) were positively associated with total depressive symptoms. Energy intake from breakfast (PR = 1.2 [1.0, 1.3], p = 0.017) was associated with a higher prevalence of high depressive symptoms. Energy intake from dinner (PR = 0.81 [0.69, 0.94], p = 0.007), later timing of the first eating episode (PR = 0.83 [0.70, 0.99], p = 0.034) and first energy quartile (PR = 0.84 [0.70, 1.0], p = 0.048), were associated with a lower prevalence of high depressive symptoms.

**Conclusion:** These findings extend prior chrononutrition-depression literature to the prenatal period, implicating eating frequency, energy intake, and meal energy timing and distribution in depressive symptomatology during pregnancy, warranting further longitudinal investigation.

**Key Messages:**

- Eating frequency, snacking, nighttime snacking, and longer eating windows were positively associated with increased prevalence of depressive symptomatology within the prenatal period. Each of these results align with previous literature in general adult or postpartum populations and extend to prenatal populations specifically. These results suggest the role of circadian misalignment (potentially via mechanisms including nocturnal cortisol response, serotonin and dopamine dysregulation, systemic inflammation, and delayed melatonin production) in prenatal psychopathology. Importantly, the potential bidirectional nature of food intake and depression cannot be overlooked.
- Total energy consumed per day, percentage of energy from breakfast and dinner, as well as later timing of initial energy intake and first caloric quartile were positively associated with prenatal depression. Additionally, particular indicators of dietary quality and macronutrient percentages (e.g., percentage of energy obtained from fat), were not significantly associated with prenatal depression. These findings call for future research to explore nuance regarding macronutrient consumption, energy intake percentages, meal timing regularity, and the possible implications of emotionally responsive eating in the relationship between food intake and prenatal depression
- The current study is exploratory in nature, being the first to explore a robust range of food intake indicators and their relationship to depressive symptomatology in the prenatal period. As prenatal depression is a strong predictor of worsening mental health during postpartum, it is important that future research test a priori hypotheses regarding food intake indicators and prenatal depression longitudinally in order to determine both temporal precedence and validate the associations found within our study. If validated, multiple domains of food intake may potentially provide modifiable behaviors that can protect against depression during the prenatal period.

## Introduction

Rates of depression and anxiety symptoms reach up to 20% in US pregnant populations (Dennis et al., 2017; Woody et al., 2017). These high rates are concerning as they contribute to poor household functioning, relationship difficulties, lower quality of life, suicidal ideation, and impaired child attachment post-delivery (Slomian et al., 2019). Several studies indicate that the negative outcomes of perinatal depression are preventable (Kobylski et al., 2023; Wahlbeck & Mäkinen, 2008), necessitating the identification of risk factors and behaviors to inform prevention and treatment efforts. Although well-established associations exist between food intake and depression in general populations (Guentcheva et al., 2020; Kabasakal-Cetin & Aydin, 2025; Kobayashi et al., 2025a; Lee & Shin, 2019), the limited amount of literature regarding prenatal depression and food intake focus on macronutrient intake and diet quality (Baskin et al., 2015; Sparling et al., 2017), largely ignoring other important domains of food intake (e.g., direct intake timing, energy intake timing, meal/snack structure, energy intake distribution across meals).

Chrononutrition refers to the amount, regularity, and timing (ART) of food intake. More specifically, chrononutrition encapsulates one of four health behaviors (i.e., eating, sleep, physical activity, light/dark exposure) that help entrain the circadian timing system (CST), a biological clock that synchronizes biological processes within the 24-hour day cycle (Conlon et al., 2023). The extant literature identifies several impacts of chrononutrition on facets of cardiovascular health (e.g., obesity, gestational weight gain, type 2 diabetes, CVD; Almoosawi et al., 2019; Chellappa et al., 2019; Hawkins et al., 2025; Katsi et al., 2022; Mason et al., 2020), with emerging literature beginning to investigate the role of chrononutrition indicators in depleted serotonin production, heightened cortisol levels, and disrupted circadian alignment, each leading to consequences including increased risk of depression and anxiety (Tjakradidjaja, 2024).

Moreover, there is current evidence linking specific isolated indicators (e.g., nighttime snacking, eating window, and snack frequency) to postpartum depression (Kobayashi et al., 2025a), consistent with evidence seen in general adult populations (Guentcheva et al., 2020; Kabasakal-Cetin & Aydin, 2025; Lee & Shin, 2019). Further studies have found late night eating to be associated with delayed melatonin onset and increased systemic inflammation, each contributing to decreased emotional stability (Kim et al., 2025). Although this suggests circadian-aligned meal timing as a noninvasive approach that may bolster both metabolic and emotional health (Kim et al., 2025), a multi-indicator examination of food intake and depression is required to capture the complex nature of dietary patterns and mental health within the perinatal period.

Despite recent interest in chrononutrition and mental health, several critical knowledge gaps remain. 1) Existing literature pertaining to chrononutrition and mental health have primarily focused on child, adolescent, and the general adult populations, largely ignoring perinatal populations (Smith et al., 2021). 2) The limited literature that does exist regarding food intake in perinatal populations focus on the postpartum period and predominately examine dietary quality, nutrient composition (e.g., Mediterranean diet adherence, omega-3 fatty acids, B vitamins, vitamin D; Baskin et al., 2015; Sparling et al., 2017), or specific individual timing indicators (i.e., nighttime snacking, breakfast skipping) within postpartum populations (Kobayashi et al., 2025a). As depression in pregnancy is a strong predictor of worsening or maintained depression during the postpartum period, understanding the role of food intake during the prenatal period is critical (O’Hara et al., 1984). Additionally, with the current understanding that chrononutrition and eating behaviors are more nuanced than macronutrient intake or the timing of isolated eating episodes alone, a broad range of potential food intake indicators that drive prenatal depression, relative to one another, is left unknown.

Thus, this study aims to address research gaps by exploring the relationship between individual mid-pregnancy food intake indicators within five general domains (i.e., intake timing, energy intake distribution across meals, energy intake timing, meal/snack frequency, overall diet quality) and prenatal depressive symptomatology in order to provide a more comprehensive view of dietary behavior and mental health in pregnancy.

## Methods

### Study Design

This is a secondary data analysis of the Comparison of Two Screening Strategies for Gestational Diabetes (GDM2) trial. This trial was a parallel-arm, single-center, comparative effectiveness trial that compared perinatal outcomes within pregnant individuals that randomly received either a Carpenter-Coustan gestational diabetes (GDM) or the International Associations of Diabetes and Pregnancy Study Groups (IADPSG) screening approach (Abebe et al., 2017; Davis et al., 2021). Participants were recruited from 10 different obstetrics clinics that were affiliated with the University of Pittsburgh Medical Center Magee-Women’s Hospital from June 2015 to February 2019. The study design and procedure were approved by The University of Pittsburgh Institutional Review Board in accordance with the Declaration of Helsinki. Participants provided written informed consent before enrollment. Oversight was provided by the data and safety monitoring board with the Clinical and Translation Science Institute at the University of Pittsburgh. The GDM2 trial was registered at clinicaltrials.gov (NCT02309138).

Detailed information regarding the protocol for the source dataset can be found in the parent study (Abebe et al., 2017; Davis et al., 2021). Participants were recruited between 18 to 28 weeks of gestation, before routine GDM screening. Participants were between 18 and 45 years and had a singleton pregnancy at the time of screening. Exclusion criteria included: (1) diabetes diagnosed <24 weeks gestation, (2) preexisting type 1 or 2 diabetes mellitus, (3) corticosteroid use in the past 30 days, (5) hypertension requiring medications, (6) anticipated a preterm delivery due to fetal or maternal indications prior to 34 weeks’ gestation, (7) severe liver disease, (8) advanced HIV, or (9) gastric bypass surgery or other conditions that prevented the participant from being administered a glucola solution. 921 participants were enrolled within GDM2. This secondary data analysis was limited to participants with 24-hour food recall and depressive symptoms data at base (described in detail below).

### Diet assessment and food intake indicators

Food intake was measured at baseline (visit 1) between 24-28 weeks’ gestation using two 24-hour dietary recalls on random nonconsecutive weekdays. The Automated Self-Administered 24-hour (ASA24®) Dietary Assessment Tool was used to collect data, which creates detailed reports on food group consumption and nutrient intake (Subar et al., 2012). An average of the two assessments was used to calculate food intake indicators within five general domains (see Table 1). An eating episode was defined as the consumption of a calorie-containing meal or beverage. Intake categories were a subjective self-report measure in which participants labeled their eating episodes as breakfast, brunch, lunch, dinner, or a snack. We define breakfast, lunch, and dinner as the first, middle, and final main meals of the day, respectively. Brunch is defined as a late-morning meal eaten in addition to or in lieu or breakfast and lunch, while snacks are defined as any energy intake that was not a main meal.

**Table 1.**
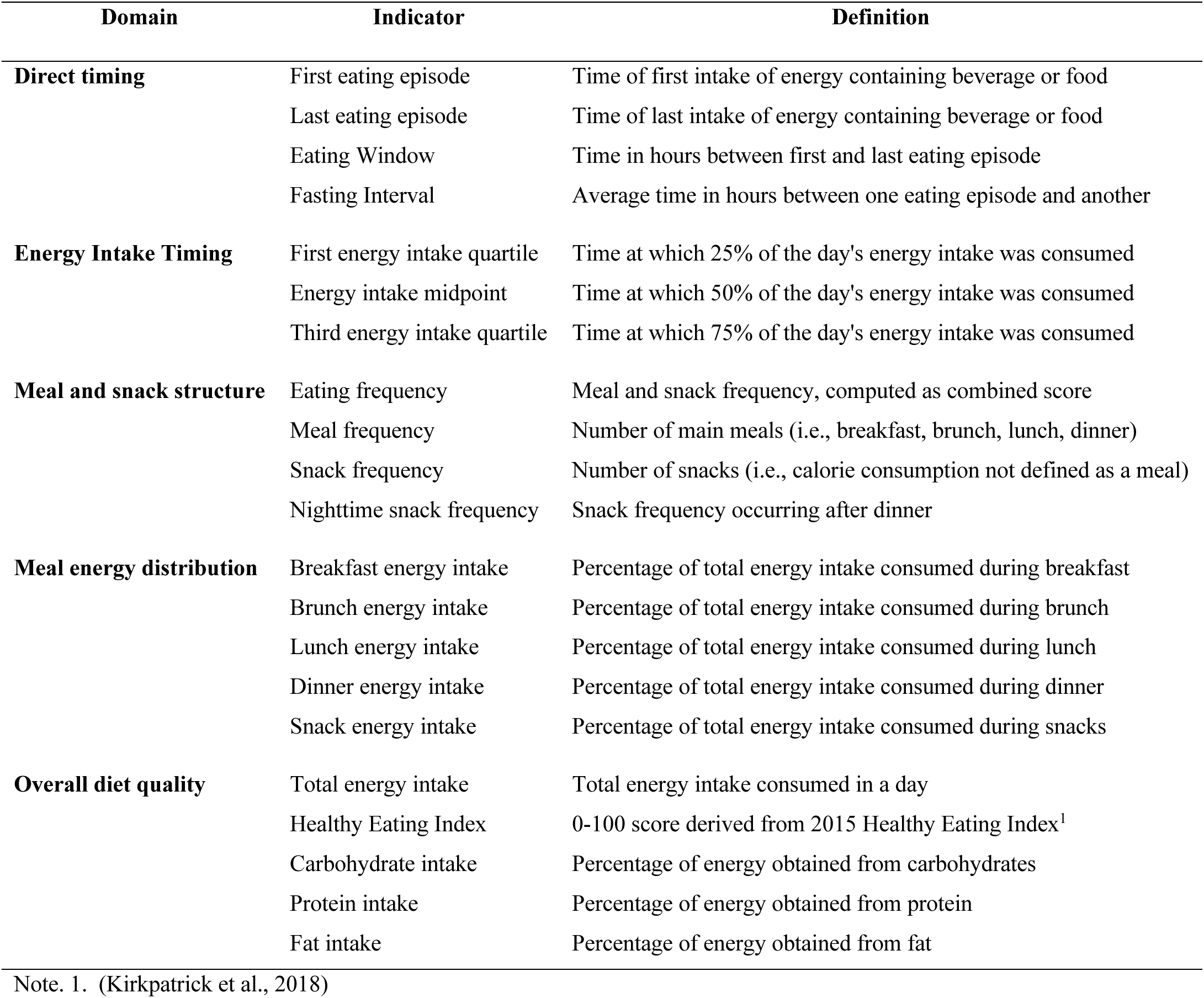
Food intake indicators by domain.

| **Domain** | **Indicator** | **Definition** |
| --- | --- | --- |
| **Direct timing** | First eating episode | Time of first intake of energy containing beverage or food |
|  | Last eating episode | Time of last intake of energy containing beverage or food |
|  | Eating Window | Time in hours between first and last eating episode |
|  | Fasting Interval | Average time in hours between one eating episode and another |
| **Energy Intake Timing** | First energy intake quartile | Time at which 25% of the day's energy intake was consumed |
|  | Energy intake midpoint | Time at which 50% of the day's energy intake was consumed |
|  | Third energy intake quartile | Time at which 75% of the day's energy intake was consumed |
| **Meal and snack structure** | Eating frequency | Meal and snack frequency, computed as combined score |
|  | Meal frequency | Number of main meals (i.e., breakfast, brunch, lunch, dinner) |
|  | Snack frequency | Number of snacks (i.e., calorie consumption not defined as a meal) |
|  | Nighttime snack frequency | Snack frequency occurring after dinner |
| **Meal energy distribution** | Breakfast energy intake | Percentage of total energy intake consumed during breakfast |
|  | Brunch energy intake | Percentage of total energy intake consumed during brunch |
|  | Lunch energy intake | Percentage of total energy intake consumed during lunch |
|  | Dinner energy intake | Percentage of total energy intake consumed during dinner |
|  | Snack energy intake | Percentage of total energy intake consumed during snacks |
| **Overall diet quality** | Total energy intake | Total energy intake consumed in a day |
|  | Healthy Eating Index | 0-100 score derived from 2015 Healthy Eating Index<sup>1</sup> |
|  | Carbohydrate intake | Percentage of energy obtained from carbohydrates |
|  | Protein intake | Percentage of energy obtained from protein |
|  | Fat intake | Percentage of energy obtained from fat |

Note. 1. (Kirkpatrick et al., 2018)

### Depression

Depression was the main outcome and was measured at baseline (visit 1) between 24-28 weeks’ gestation using the Edinburgh Postnatal Depression Scale (EPDS) (Cox et al., 1987). The EPDS is a 10-item self-report questionnaire with scores ranging from 0 to 30, with higher scores corresponding with greater depressive symptom severity. Depression score was log-transformed to manage right-sided skewness. Depression was also dichotomized based on clinical cutoffs moderate (EPDS score of ≥10) and high depressive symptom severity (EPDS score ≥13) (Cox et al., 1987; Harris et al., 1989; Murray & Carothers, 1990). Participants that scored a 13 or above on the EPDS were provided supportive resources for depression. The EPDS has acceptable specificity (78%) and sensitivity (86%) for predicting depression (Cox et al., 1987) and is well validated for use in pregnant populations (Bergink et al., 2011; Zeng et al., 2025).

### Descriptive Characteristics

Demographic characteristics included self-reported age and race (i.e., American Indian/Alaska Native, Asian, Black, Native Hawaiian or Pacific Islander, White, or more than one race). Additionally, participants were asked if they were Latino or Hispanic. Due to small sample sizes, participants were grouped according to the United States census as Black, White, Asian, and “all other racial identities”.

Baseline covariates included psychosocial, and behavioral risk factors for depression, as well as factors that could dictate food intake patterns. Participants self-reported employment status (i.e., part-time, fulltime, or not working), marital status (i.e., single/divorced or married/living with a partner, widowed), education (i.e., high school/GED or less, some college/vocational school, college degree, beyond college), number of adults living within the household, income (i.e., ≤ $30,000, $31,000 - 60,000, ≥ $61,000), current smoking status (i.e., cigarettes, e-cigarettes, or cigars per day), and current alcohol consumption (i.e., yes, no). Self-reported current health status (e.g., poor, fair, good, excellent) was grouped by poor/fair and good/excellent.

Physical activity was self-reported and measured using the Godin Leisure-Time Exercise Questionnaire, a tool that assesses weekly mild, moderate, and strenuous exercise during leisure time (Amireault & Godin, 2015). A Leisure Score Index was computed by summing the frequency of mild, moderate, and strenuous activities and multiplying the type of activity by its respective metabolic equivalent task value. Insomnia symptom frequency was assessed through self-reporting on how often participants experienced trouble staying asleep, trouble falling asleep, waking several times per night, and waking after their usual amount of sleep feeling tired or worn out in the past month. Scores for each self-report question ranged from 0 (not at all) to 5 (22 to 31 days). Total insomnia symptom frequency was identified by summing scores for each question and grouping them into tertiles (i.e., 0–6, 7–13, and 14–20). Higher scores indicated more frequent insomnia symptoms. Current health status was self-reported as poor, fair, good, or excellent. Participant responses were categorized into two groups: poor/fair and good/excellent. Perceived stress was measured using the Perceived Stress Scale (Cohen et al., 1983), a 14-item scale used to assess the frequency of stress experienced by participants in the past month. Scores range from 0 to 56, with higher scores indicating more perceived stress. Test-retest correlations ranged between 0.55 and 0.85 within the general population (Cohen et al., 1983).

### Statistical Analysis

For the primary aim, linear regression was run to examine the association between mid-pregnancy food indicators within five general domains (e.g., direct intake timing, energy intake timing, meal/snack structure, energy intake distribution across meals, overall diet quality) and total prenatal depression scores (outcome). All food intake indicators were run within separate models. Relevant covariates that could potentially impact either exposure or outcome variables were added into the main analysis. Covariates were identified based on the conceptual expertise of the research team and included age, income, education, marital status, working status, health status, perceived stress, insomnia frequency, physical activity, and both current alcohol and smoking use. Diabetes status and the treatment assignment were also included in the models to preserve the study design of the parent study.

For the secondary aim, Poisson regression was employed within separate adjusted models to examine the association between food intake indicators and moderate and high prenatal depression. Poisson regression with a sandwich estimator was chosen to produce prevalence ratios (PR) for both ease of interpretation and to avoid overestimation of prevalence that can occur when using logistic regression (Barros & Hirakata, 2003).

Although analyzing multiple indicators increases the risk of Type 1 error, our study is exploratory in nature and thus corrections were not employed (Hooper, 2025). A post-hoc sensitivity analysis was run in the Poisson model for findings that misaligned with hypothesized directions to examine potential impacts of diet quality (specifically the healthy eating index) on initial findings. For all models, numeric variables were scaled to better compare coefficients. Missing data was accounted for using case wise deletion in both primary and secondary analysis. An alpha level of p = 0.05 was deemed statistically significant for hypothesis testing. All analyses were run in R/RStudio 4.5.1 (R Core Team, 2024).

## Results

### Initial vs. final sample comparison

Of the 921 initial participants enrolled in the GDM2 study, participants with missing food intake and depression variables (n = 203) were excluded in the present analysis, leaving 718 participants (See Supplementary Material). Participants included in the analyses were older, of higher socioeconomic status and educational level, and were more likely to be currently married and employed compared with excluded participants. They also had fewer depressive symptom, perceived stress, and were less likely to smoke (See Supplementary Material). Within the included sample, individuals in the high depression group reported lower socioeconomic status, lower educational attainment, and lower marriage rates. Additionally, the high depression group was more racially and ethnically diverse, had lower employment rates, and lower self-reported health ratings. Subsequently, the high depression group endorsed more insomnia symptoms, slightly lower physical activity levels, and more alcohol/tobacco use. All groups had similar distributions of dietary quality, total energy intake per day, and macronutrient consumption (e.g., carbohydrate, protein, fat). Detailed sample characteristics by depression category (e.g., low, moderate, high) can be found in Table 2.

**Table 2.**
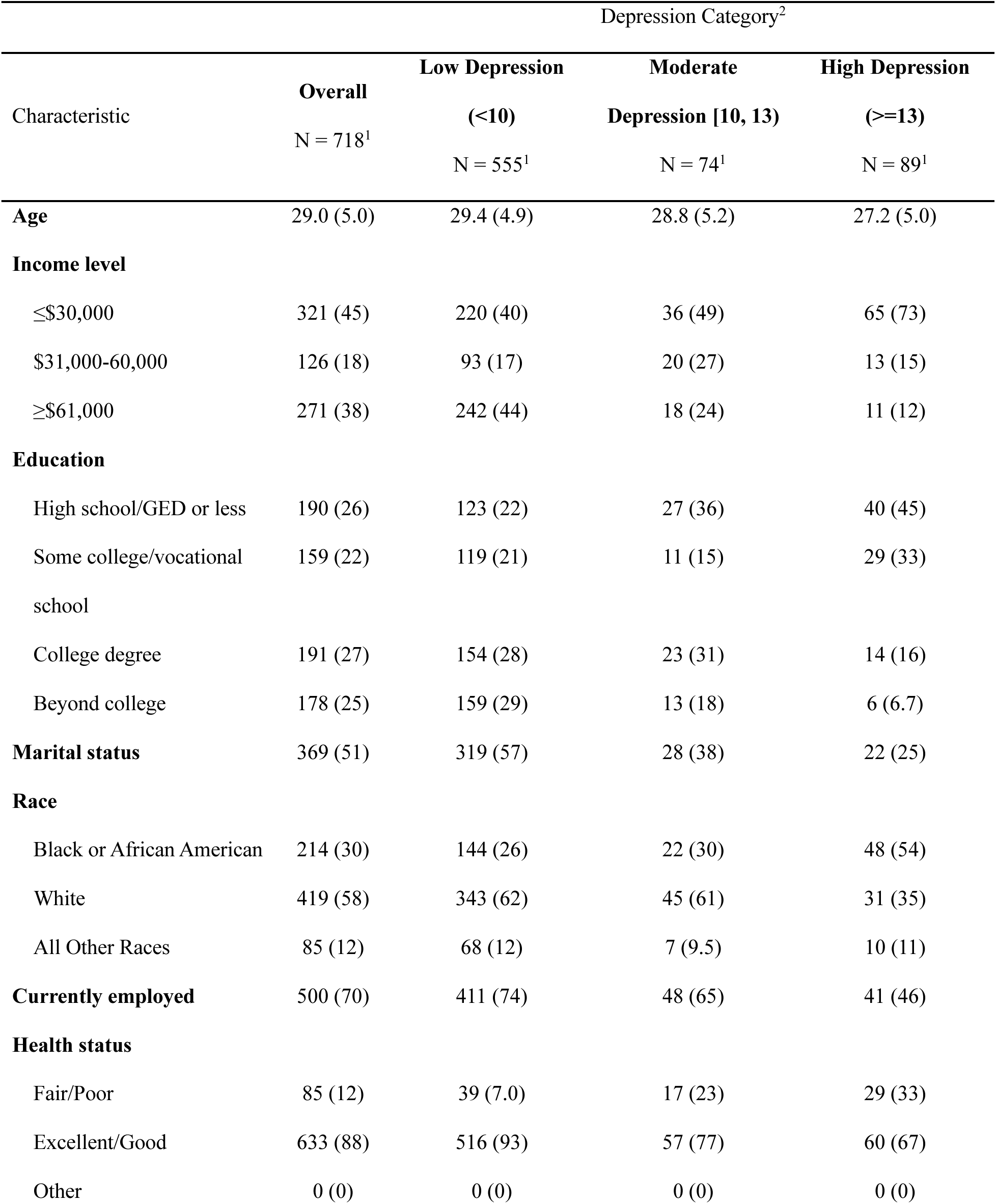
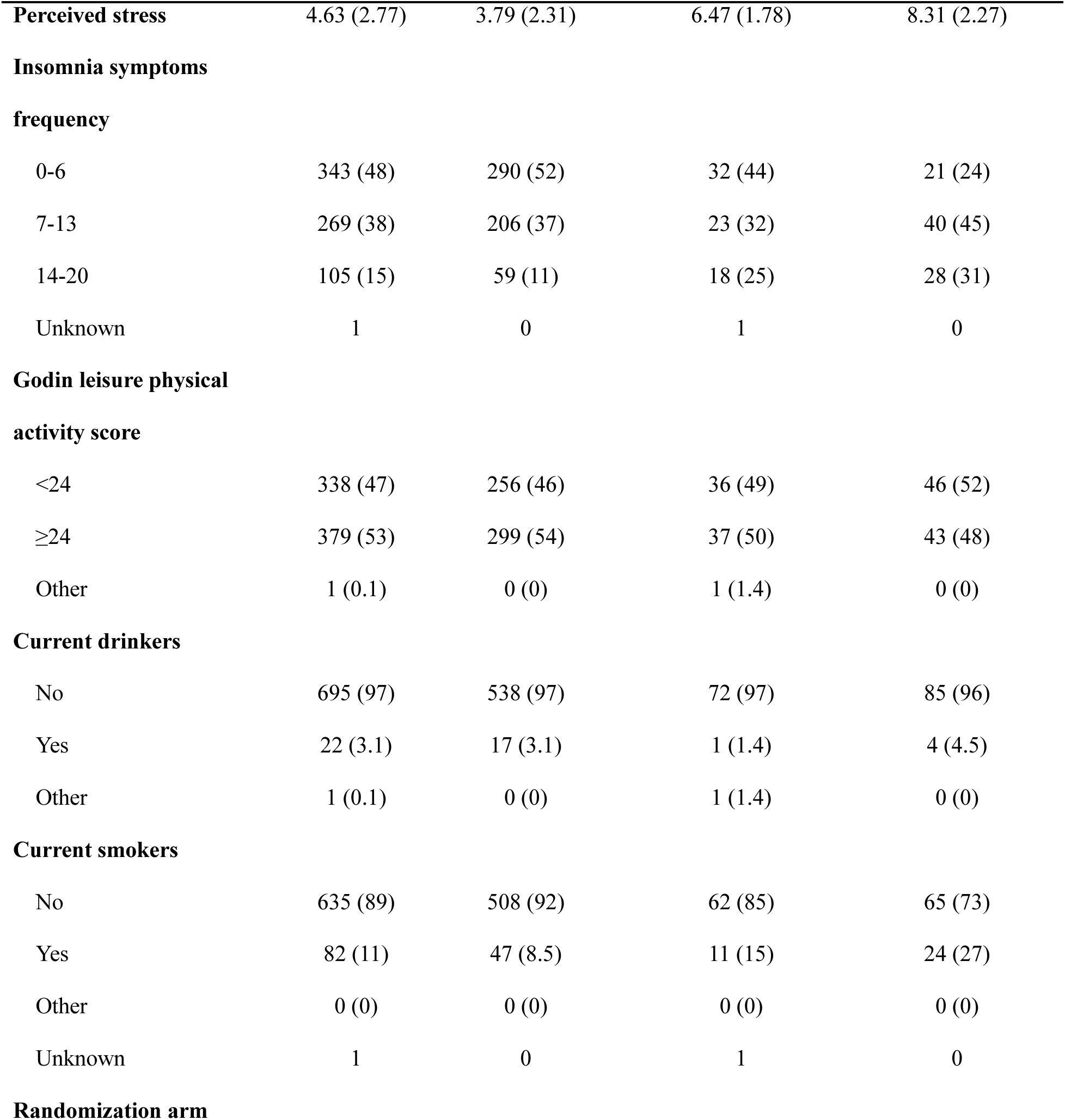
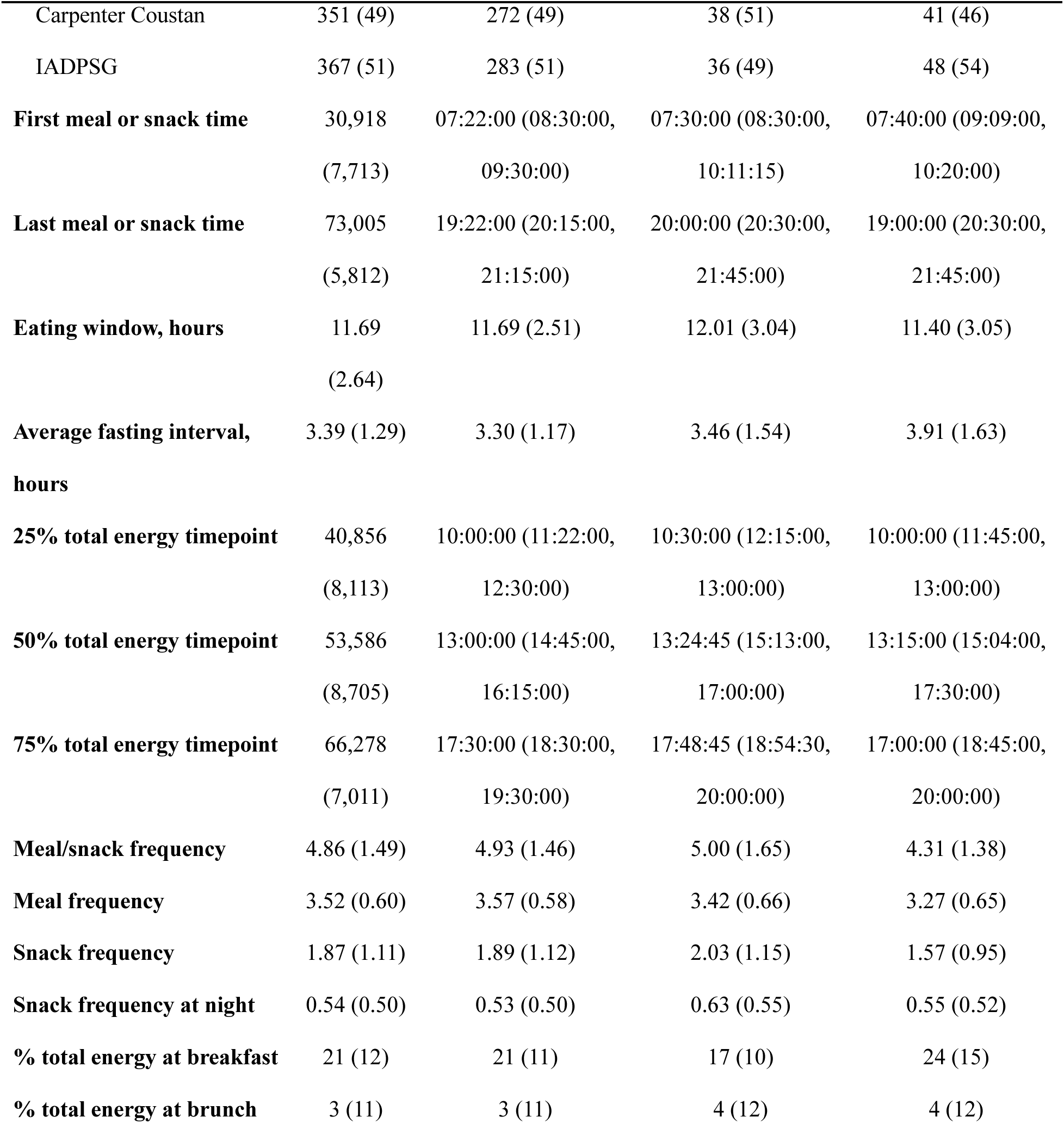
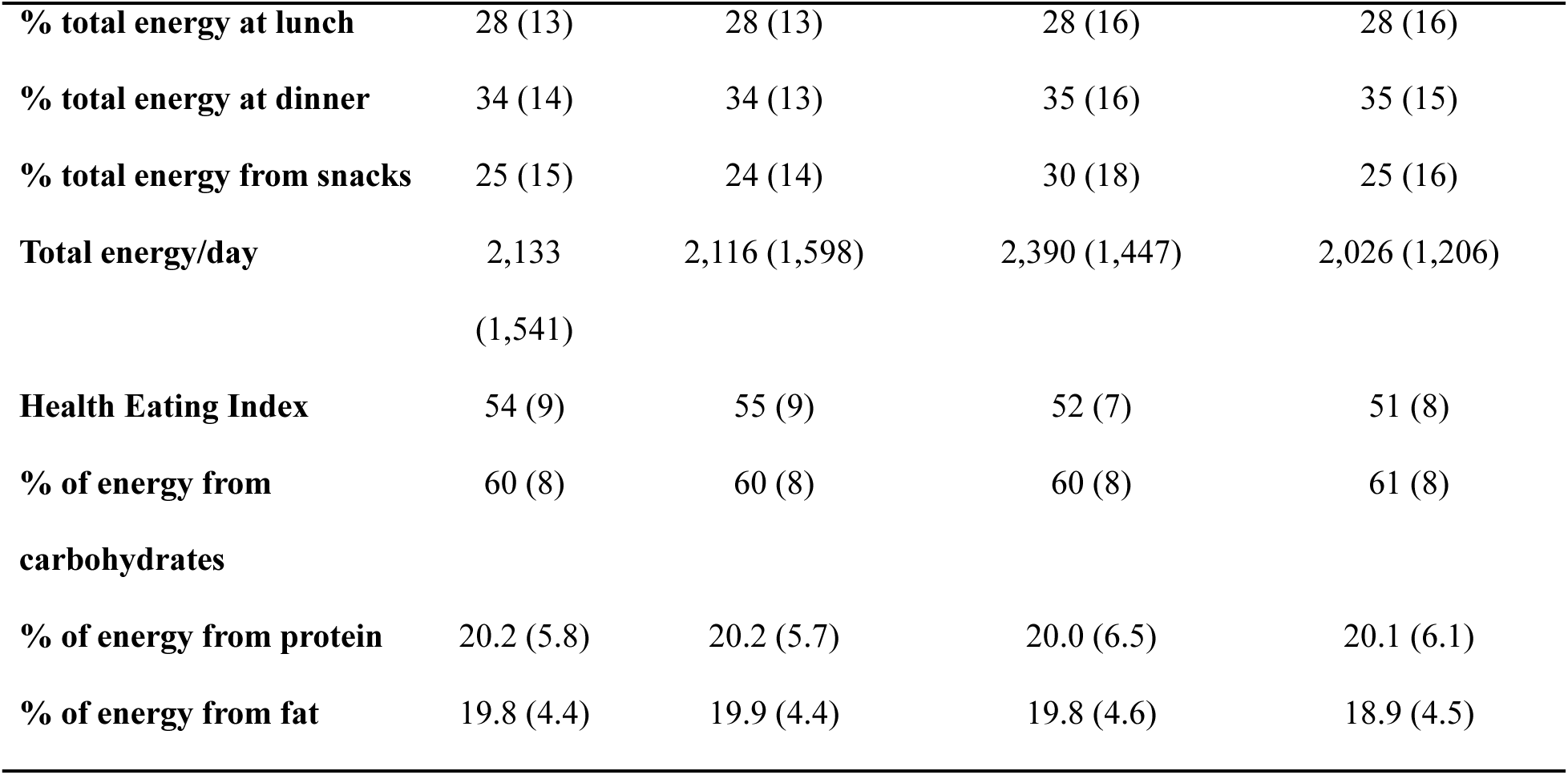
Descriptive characteristics by depression symptom severity.

### Main analysis

In linear regression models, there was a positive relationship between eating frequency and log-depressive symptoms when controlling for covariates (B = 0.08 [0.02, 0.14], p = 0.009), indicating an 8% multiplicative increase in average depression score for each standard deviation increase in eating episodes. Similarly, there was a positive relationship between log-depressive symptoms and snack frequency (B = 0.06 [0.00, 0.12], p = 0.04), as well as nighttime snacking frequency (B = 0.6 [0.00, 0.11], p = 0.041). There was also a significant positive association for total daily energy intake (B = 0.06 [0.01, 0.12], p = 0.031). Regression results for log-depressive symptoms can be visualized in Figure 1.

**Figure 1.**
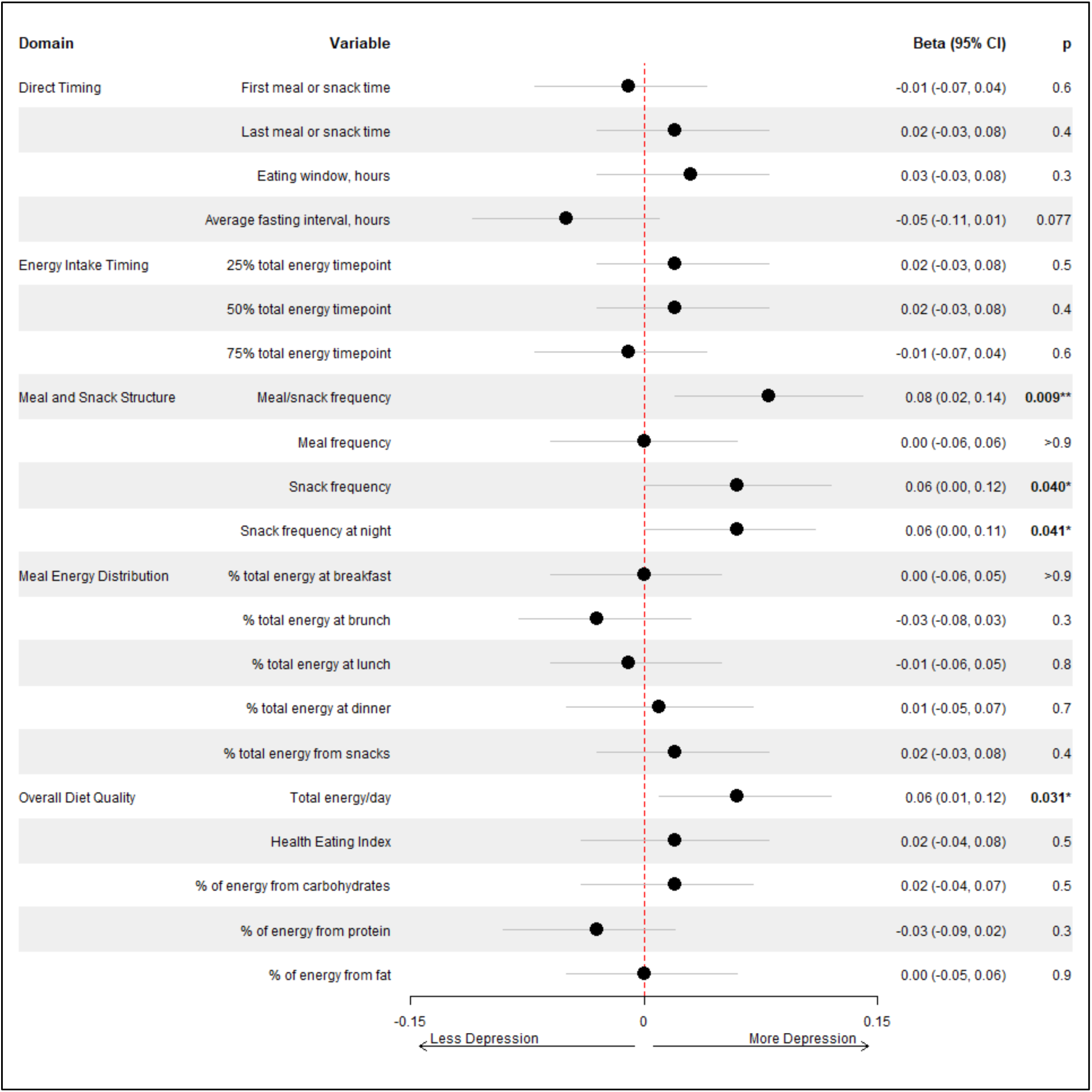
The adjusted association between food intake indicators and log-transformed prenatal depressive symptoms.

In secondary analysis, there was a positive relation between total energy intake and moderate depression (PR = 1.1 [1.0, 1.3], p = 0.042) after adjusting for covariates, indicating a 10% higher prevalence in moderate depression for every standard deviation increase in energy intake. There was also a significant positive relation between eating window and moderate depression (PR = 1.1 [1.0, 1.3], p = 0.034). Interestingly, there were several findings that suggested a negative relationship between later food timing and high depression. There was a significant negative relation between the time of the first eating period and high depression (PR = 0.83 [0.70, 0.99], p = 0.034), indicating a 17% lower prevalence of high depression for each standard deviation increase in first meal or snack time. In addition, there was a negative relation between the time 25% total energy was consumed and high depression (PR = 0.84 [0.70, 1.0], p = 0.048). There were also negative relationships between high depression and percentage of energy from breakfast (PR = 1.2 [1.0, 1.3], p = 0.017) and percentage of total energy at dinner (PR = 0.81 [0.69, 0.94], p = 0.007). Post-hoc sensitivity analyses in which diet quality was added into the model did not attenuate these results. Robust Poisson regression analyses showing main effects for moderate and greater depressive symptom severity can be visualized in Figures 2 and 3, respectively.

**Figure 2.**
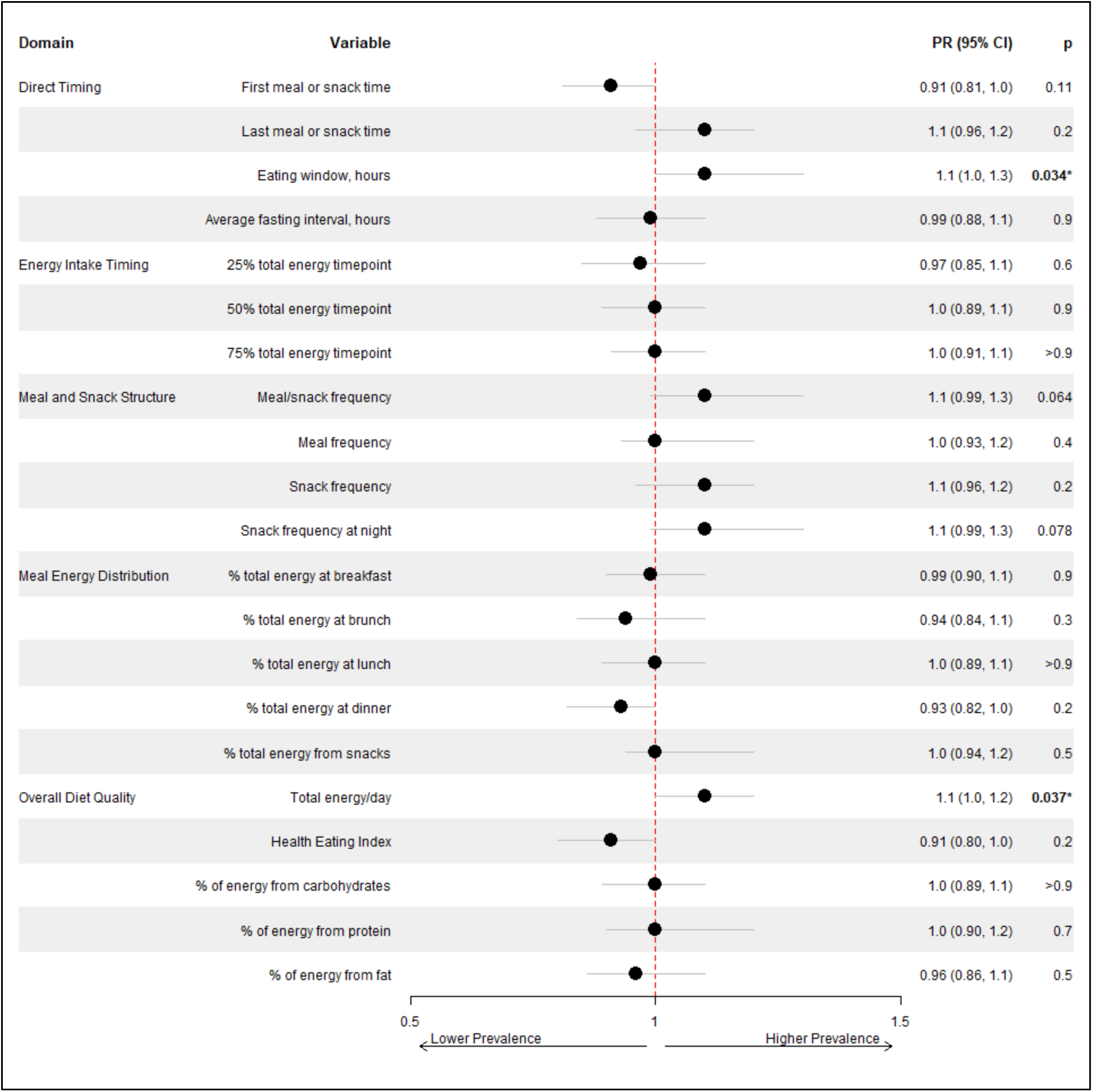
The adjusted association between food intake indicators and moderate prenatal depressive symptoms.

**Figure 3.**
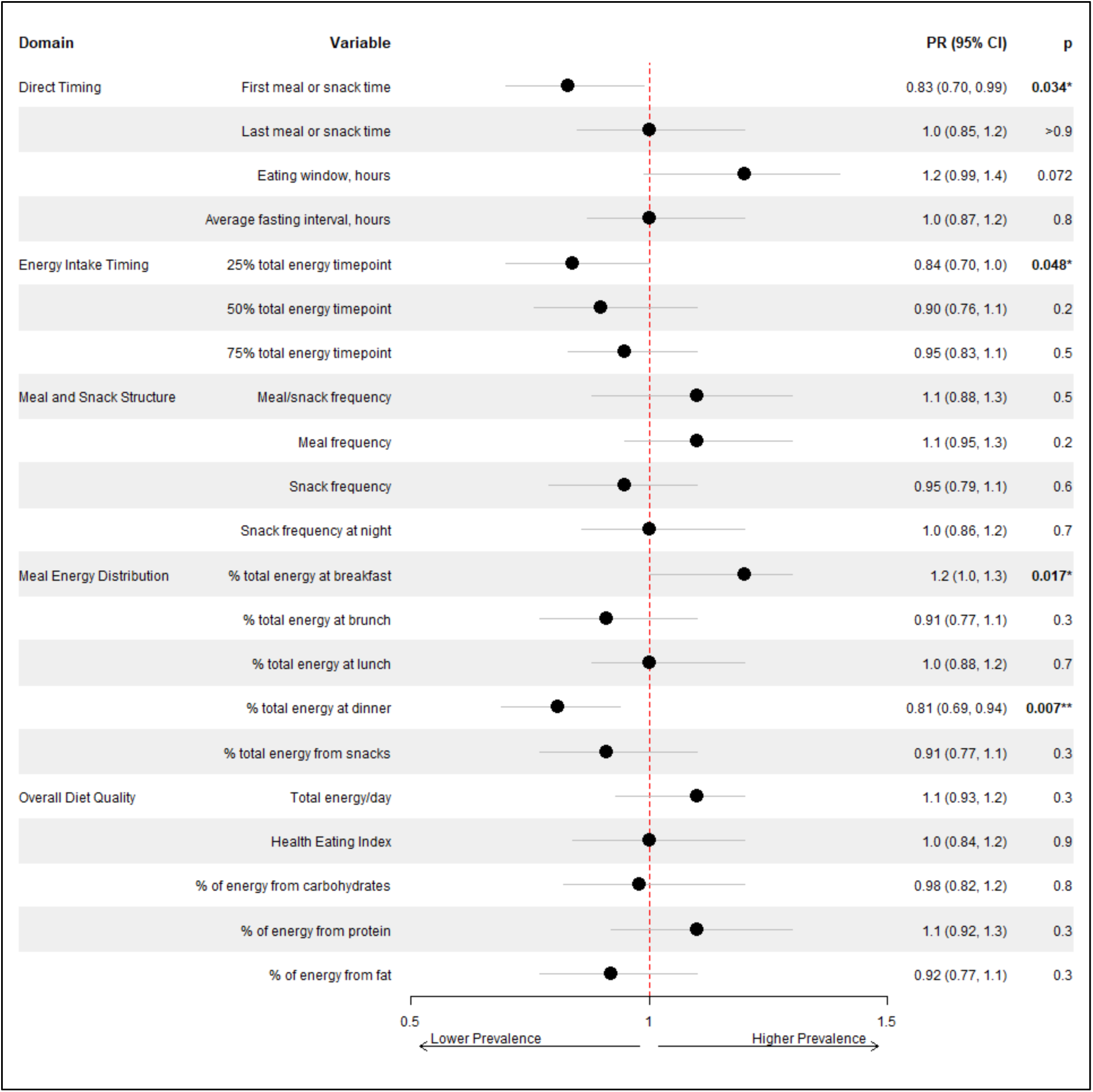
The adjusted association between food intake indicators and high prenatal depressive symptoms.

## Discussion

This study found positive associations between eating frequency, nighttime snacking frequency, and total depressive symptomatology, consistent with previous links between late night eating episodes and depressive symptoms in both postpartum (Kobayashi et al., 2025b) and general populations. Associations between eating frequency and nighttime snacking frequency may implicate circadian-misaligned eating in prenatal depression, which has previously been shown to impact numerous biological mechanisms related to sleep (i.e., nocturnal cortisol response, serotonin and dopamine dysregulation, systemic inflammation, and delayed melatonin production; Kim et al., 2025). Each of these proposed mechanisms provide potential rationale for the associations regarding heightened depressive symptomatology within this sample. Additionally, snacking frequency was found to be positively associated with prenatal depressive symptomatology, supporting previous findings in general populations (Furihata et al., 2018). A 2014 study by Camilleri et al. suggests associations between depression, energy dense snacking, and emotionally responsive eating in women, a particular facet that was not explored in the present analysis. Within our study, nighttime snacking frequency does not provide insight regarding calorie density or macronutrient content of post-dinner food intake. As such, the interactions between night-time snacking, emotionally driven eating behavior, caloric and macronutrient intake in pregnancy warrant further investigation. Due to the cross-sectional nature of this analysis, the possibility that increased depressive symptomatology may contribute to increased food intake indicators must also be considered.

In addition to meal and snack structure, the current study found positive associations regarding total daily energy consumed and continuous prenatal depressive symptomatology. While previous literature does not suggest links regarding particular daily caloric thresholds and postpartum depression, excessive gestational weight gain and emotional eating are known to be strong predictors of postpartum depression (Wu et al., 2023) and should be considered in future literature when examining total diet quality and prenatal depressive symptomatology.

Further, energy intake, weight, and mood are largely considered bidirectional and cyclical in nature. Although intake of energy dense foods may offer transient mood improvement, long term adherence to an energy dense and high-fat diet has been shown to be associated with weight gain and subsequent negative affective states, creating a self-reinforcing drive to seek further mood improvement through high-calorie food intake (Calcaterra et al., 2024; Singh, 2014). Our findings generate further questions, as we did not observe significant findings nor meaningful effect sizes regarding the percentage of energy obtained from fat. Moreover, with previous literature finding antenatal depression as a predictor for lower maternal diet quality, our null findings regarding several indicators of diet quality and prenatal depressive symptomatology warrant further, longitudinal investigation. It should be noted that the little variability of diet quality within this sample may have decreased the power of our analysis, making it difficult to detect effects. Additionally, the use of a single point dietary recall assessment may not provide the sensitivity needed in order to comprehensively assess the link between diet quality and prenatal depression and is a limitation that should be considered in future studies.

Contrary to previous literature, results from the main analysis showed positive associations between higher percentage of energy obtained through breakfast and high prenatal depressive symptomatology. These results do not align with previous research finding positive associations between breakfast skipping and depression in general populations (Li et al., 2024; Zahedi et al., 2022), with additional literature showing decreased depression risk in prenatal individuals when 5% of total energy obtained from dinner was isocalorically replaced with breakfast obtained energy (Jiang et al., 2026). However, our findings do not account for the macronutrient composition of breakfast consumption. Although recent literature links breakfast quality and depressive symptomatology, implicating the impact of quality breakfast on metabolic and appetite regulation, insulin levels, and blood glucose (Sun & Wu, 2025), our results remained unchanged in both effect size and significance after exploring this association with a diet quality indicator introduced into the model during a post-hoc sensitivity analysis. Additionally, sleep timing was not considered in the present analysis and should be considered in future research, as it may provide a more robust picture in the context of depression (e.g., depressed individuals may sleep later in the day and report lunch as their first meal, having impacts on increased energy consumption for participant defined “breakfast”).

In addition to the potential implications of macronutrient consumption and sleep timing, our results cannot determine the direction of the relationship between prenatal depression and energy obtained through breakfast. Although previous literature has found emotionally responsive eating to be associated with depressive symptomatology (Konttinen et al., 2010; van Strien et al., 2016), it also suggests close associations with night eating syndrome and evening types (Garaulet et al., 2023; Nolan & Geliebter, 2012; Shillito et al., 2018). As our findings also suggest that higher energy intake during dinner is associated with lower prevalence of high prenatal depressive symptomatology, these data inadequately explain why positive associations regarding high depressive symptomatology were observed with higher breakfast energy intake and not later meals. Thus, it is critical that future research conduct longitudinal studies that test a priori hypotheses regarding the strength and direction of association regarding prenatal depression and the percentage of energy obtained through breakfast.

Additionally, positive associations were found between later timing of first eating episode, first energy intake quartile, and decreased prevalence of high prenatal depressive symptomatology. This does not align with previous research that consistently shows delayed first eating episodes to be associated with poorer physical and mental health outcomes (Dashti et al., 2025). However, we also found that longer eating windows were associated with moderate prenatal depression, a result that is supported by previous literature finding eating windows greater than twelve hours to be positively associated with depression occurrence (Li et al., 2024). Although previous literature does find associations between delayed or skipped breakfast and depression risk, more current research suggests that meal timing irregularity, not breakfast skipping, is an independent predictor of postpartum depression (Tahara et al., 2026). This result in the context of our significant finding regarding longer eating windows and moderate prenatal depressive symptomatology may better explain the found associations, although the possibility of Type I error is still present. As our analysis did not consider the consistency or regularity of participant mealtimes, these results warrant further investigation into the topic when examining food intake timing and prenatal depressive symptomatology.

The significant associations we observed between various domains of food intake, including direct intake timing, energy intake timing, meal and snack structure, overall diet quality, meal energy distribution, and prenatal depressive symptomatology suggest that food intake is a relevant for mood disorders not only in the postpartum period but also during pregnancy. Previous literature suggests that prenatal depression accounts for half of the variance seen in postpartum depressive symptomatology (O’Hara et al., 1984). Considering this, intentional focus should be shifted towards examining, preventing, and treating depression prenatally to buffer severe progression of symptoms after birth, a time that is crucial for a pregnant individual’s physical healing and infant bonding (Johnson, 2013; Schwartz et al., 2025). The findings of this study provide insight into modifiable behaviors linking mid-pregnancy dietary patterns to prenatal depression risk and raise questions for future examination.

### Limitations and Future Directions

Although the current study provides a robust foundation to guide future discovery within a diverse and representative sample, limitations must be discussed. First, due to the cross-sectional nature of the analysis, we cannot determine whether 1) food intake indicators caused increased depression, 2) increased depression caused food intake misalignment, or 3) that this relationship is cyclical, with each factor reinforcing the other. Additionally, both primary predictors and outcome measures were obtained through self-report, potentially contributing to shared method variance that might account for portions of the observed associations. Although the EPDS is a well validated scale and is widely used in studying depression in perinatal populations, using clinical assessments for depression may enhance the robustness of the findings. Similarly, we assessed food intake using a dietary recall within one time point in mid pregnancy, and future research may benefit from assessing dietary intake at various points within pregnancy for a more comprehensive understanding of food intake patterns. It must also be mentioned that only insomnia symptoms were monitored. A multidimensional assessment of sleep (e.g., timing, regularity, duration, satisfaction, and efficiency) may provide a more robust understanding of how the synergistic nature of circadian-driven behaviors relate to mental health in pregnancy.

Lastly, the examination of several indicators increases the likelihood of Type I error, and corrections were not employed due to the exploratory nature of our study (Hooper, 2025). Our findings should be interpreted as steppingstone to guide future research and generate hypotheses rather than definitive, or causal findings. Future research should consider these implications when intending to extend or validate the relationship between food intake and prenatal depression.

## Conclusion

The current study found that eating frequency, snacking, nighttime snacking, and longer eating windows were positively associated with depression within the prenatal period, suggesting the role of circadian misalignment in prenatal psychopathology. Further, our study raises questions regarding the implications of total energy consumed per day, percentage of energy from breakfast and dinner, as well as later timing of initial energy intake and first caloric quartile and prenatal depression. These findings call for future research to explore nuance regarding macronutrient consumption, energy intake percentages, meal timing regularity, and the possible implications of emotionally responsive eating in the relationship between food intake and prenatal depression.

## Author Contributor Statement

CMP, DP, and MSH performed the research and MSH designed the research study. KA and ED contributed essential reagents and data. MSH, NK, DP, and CMP analyzed the data. CMP and DP wrote the paper. DD, CMS, TC, DM, PC, HS, NK, and MHS consulted on methodology, interpretation of results, and contributed to revising the manuscript. All authors have read and approved the final manuscript.

## Data Availability Statement

Data can be made available upon a reasonable request and data use agreement with the study principal investigator.

