## Supplemental Table 1 for "Chrononutrition and prenatal mental health: The relationship between food intake indicators and prenatal depressive symptomatology"

**Table S1.** Characteristics of included/excluded participants

|  | Included in Analysis | |  |
| --- | --- | --- | --- |
| Characteristic | **No**  N = 203^1^  N (%) or mean ± SD | **Yes**  N = 718^1^  N (%) or mean ± SD | **p-value**^23^ |
| Age | 27.0 (23.0, 32.0) | 29.0 (26.0, 32.0) | <0.001*** |
| Income level |  |  | <0.001*** |
| ≤$30,000 | 128 (63) | 321 (45 |  |
| $31,000-60,000 | 26 (13) | 126 (18) |  |
| ≥$61,000 | 49 (24%) | 271 (38) |  |
| Education |  |  | <0.001*** |
| High school/GED or less | 85 (42%) | 190 (26%) |  |
| Some college/vocational school | 54 (27%) | 159 (22%) |  |
| College degree | 37 (18%) | 191 (27%) |  |
| Beyond college | 27 (13%) | 178 (25%) |  |
| Married | 60 (30%) | 369 (51%) | <0.001*** |
| Race |  |  | 0.001** |
| Black or African American | 86 (42%) | 214 (30%) |  |
| White | 91 (45%) | 419 (58%) |  |
| All Other Races | 26 (13%) | 85 (12%) |  |
| Currently Employed | 121 (60%) | 500 (70%) | 0.009** |
| Health status |  |  | 0.4 |
| Fair/Poor | 17 (15%) | 85 (12%) |  |
| Excellent/Good | 94 (85%) | 633 (88%) |  |
| Perceived stress | 5.0 (3.0, 8.0) | 4.0 (2.0, 6.0) | 0.015* |
| Insomnia Symptoms Frequency |  |  | 0.042* |
| 0-6 | 64 (58%) | 343 (48%) |  |
| 7-13 | 28 (25%) | 269 (38%) |  |
| 14-20 | 19 (17%) | 105 (15%) |  |
| Physical Activity |  |  | 0.9 |
| <24 | 51 (46%) | 338 (47%) |  |
| ≥24 | 60 (54%) | 379 (53%) |  |
| Alcohol Use | 4 (3.6%) | 22 (3.1%) | >0.9 |
| Smoking | 32 (29%) | 82 (11%) | <0.001*** |
| Gestational diabetes | 7 (6.4%) | 73 (10%) | 0.3 |
| Randomization arm |  |  | 0.3 |
| Carpenter Coustan | 109 (54%) | 351 (49%) |  |
| IADPSG | 94 (46%) | 367 (51%) |  |
| Log depressive symptoms score | 1.87 (1.50, 2.44) | 1.70 (1.25, 2.25) | 0.006** |
| Moderate Depression | 34 (31%) | 163 (23%) | 0.088 |
| High Depression | 20 (18%) | 89 (12%) | 0.14 |
| First Eating Episode | 07:22:00 (08:30:00, 09:45:00) | 00:15:30 (11:00:00, 15:30:00) | >0.9 |
| Last Eating Episode | 19:30:00 (20:22:30, 21:24:15) | 15:30:00 (18:00:00, 21:45:00) | 0.060 |
| Eating window | 17.00 (9.25, 18.00) | 11.75 (10.50, 13.08) | 0.4 |
| Average fasting interval | 5.38 (3.77, 6.17) | 3.09 (2.57, 3.88) | 0.11 |
| First Caloric Quartile | 10:00:00 (11:30:00, 12:43:00) | 08:45:00 (12:00:00, 16:00:00) | 0.8 |
| Second Caloric Quartile | 13:00:00 (14:45:00, 16:30:00) | 12:00:00 (15:00:00, 16:00:00) | 0.3 |
| Third Caloric Quartile | 17:30:00 (18:37:00, 19:44:15) | 15:30:00 (18:00:00, 20:22:30) | 0.4 |
| Eating Frequency | 2.00 (1.00, 5.00) | 5.00 (4.00, 6.00) | 0.008** |
| Meal frequency | 1.50 (1.00, 3.00) | 3.50 (3.00, 4.00) | <0.001*** |
| Snack frequency | 0.00 (0.00, 2.00) | 2.00 (1.00, 2.50) | 0.071 |
| Nighttime Snacking Frequency | 0.00 (0.00, 0.00) | 0.50 (0.00, 1.00) | 0.093 |
| Breakfast Calories | 27 (14, 53) | 20 (14, 27) | 0.2 |
| Brunch Calories | 0 (0, 8) | 0 (0, 0) | 0.9 |
| Lunch Calories | 37 (20, 100) | 27 (20, 36) | 0.078 |
| Dinner Calories | 31 (27, 84) | 33 (25, 41) | 0.2 |
| Snack Calories | 32 (20, 37) | 24 (15, 34) | 0.3 |
| Total Calories | 1,316 (772, 2,602) | 2,004 (1,472, 2,516) | 0.4 |
| Health Eating Index | 45 (40, 51) | 55 (48, 60) | 0.002** |
| Carbohydrate Calories | 55 (49, 59) | 61 (55, 65) | 0.015* |
| Protein Calories | 22.0 (16.9, 26.0) | 19.5 (16.3, 23.3) | 0.13 |
| Fat Calories | 22.5 (20.7, 25.9) | 19.7 (16.6, 22.7) | 0.008** |
| ^1^Median (Q1, Q3); n (%) | | | |
| ^2^Welch Two Sample t-test; Pearson's Chi-squared test | | | |
| ^3^*p<0.05; **p<0.01; ***p<0.001 | | | |
