## Supplementary figures and images for "Chrononutrition and prenatal mental health: The relationship between food intake indicators and prenatal depressive symptomatology"

### Supplemental Figure 1

#### Slide 1
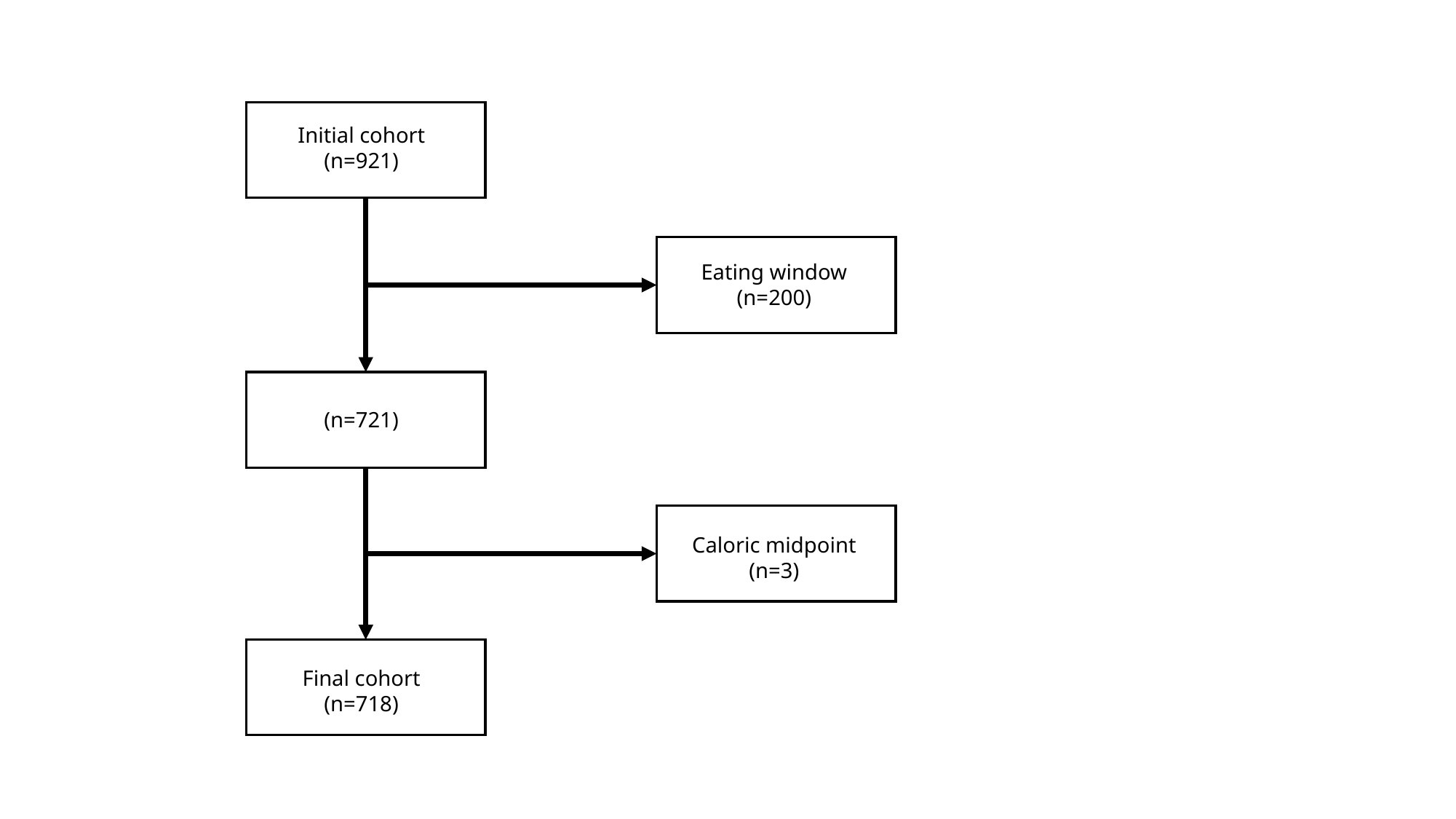

Initial cohort
(n=921)
Eating window
(n=200)
(n=721)
Caloric midpoint (n=3)
Final cohort
(n=718)
